# Significant increase of emergency department visits for heat-related emergency conditions in the United States from 2008 – 2019: a comprehensive nationwide study

**DOI:** 10.1101/2022.08.16.22278843

**Authors:** Penelope Dring, Megan Armstrong, Robin Alexander, Henry Xiang

## Abstract

**Introduction:** Exposure to high temperatures is detrimental to human health. As climate change is expected to increase the frequency of extreme heat events, as well as raise ambient temperatures, an investigation into the burden of heat-related emergency department visits is necessary to assess the human health impact of this growing public health crisis.

**Methods:** Emergency department visits were sourced from the Healthcare Cost and Utilization Project Nationwide Emergency Department Sample. This dataset collects emergency department visit information from 989 facilities that represent a 20-percent stratified sample of United States hospital-owned emergency departments. Visits were included in this study if the medical diagnosis contained an ICD-9-CM or ICD-10-CM code specific to heat-related emergency conditions. Weighted heat-related emergency department visit counts were generated to estimate the total counts for heat-related emergency department visits across the United States. Visit year and month, hospital geographic region, patient age, and sex were recorded. Incidence rates per 100,000 US population, visit counts, and visit count percent change were analyzed on both a national scale and stratified by month, region, age, and sex.

**Results:** A total of 1,007,134 weighted heat-related emergency department visits between 2008 - 2019 were included in this study. The annual incidence rate per 100,000 US population increased by an average of 5.73% (95% CI: 8.64% – 20.1%) per year across the study period, rising from 20.56 in 2008 to 30.41 in 2019. For the twelve-year period, the burden of heat-related emergency department visits was greatest in the South (51.41%). Most cases occurred in July (29.56%), with visits increasing to the greatest degree in July (19.25%, 95% CI: 20.75% – 59.26%) and March (14.36%, 95% CI: 19.53% – 48.25%).

**Conclusions:** This study found a significant increase in emergency department visits for heat-related emergency conditions across the United States from the years 2008 to 2019.

## Introduction

Scientists predict a global temperature increase exceeding 7^0^ Fahrenheit (F) above pre-industrial averages by the end of the century [1]. A sharp increase in weather-related mortality is expected across the United States as a result of this climate change [2]. As temperatures rise, episodes of extreme heat are expected to become longer, more intense, and more frequent [3,4]. The negative impact of extreme heat events on human health, as it relates to both morbidity and mortality, has been well documented [5–13].

Emergency department (ED) patient volumes rise during episodes of extreme heat, which are typically defined as periods of temperature exceeding the 95^th^ – 99^th^ percentile of region norms [5,6,14,15]. Unsurprisingly, visits for heat-related emergency conditions – a term used to encompass a spectrum of symptoms resulting from exposure to high temperatures [16] - are seen to increase by roughly 70% during these events [15,17]. Outside of extreme heat events, higher ambient temperatures are also harmful to human health [18]. Mortality risk is seen to rise by 8% for each degree (F) increase in maximum daily temperature from one day to the next [7]. Heat-related emergency conditions are also seen to increase on days with higher ambient temperature that do not meet the threshold of extreme heat, with one study citing a 393.3% excessive risk in heat-related ED visits for every 10^0^ (F) increase in temperature [19].

Almost all investigations into heat-related ED visits have restricted the time course of analysis to extreme heat events during the warm season [15,19–23]. However, the effects of climate change have necessitated a new approach to researching heat-related emergency conditions. Ambient temperatures have risen to a greater degree during the winter months as compared to summer months, and seasonal shifts have seen warmer weather arrive earlier in the Spring and linger later into the Fall [24–26]. The concern for heat-related emergency conditions is therefore no longer just limited to the warm season and should be evaluated throughout the year. Further, past investigations into heat-related ED visits have generally included narrow populations of interest, with studies focused on populations in California, Florida, and North Carolina [17,19,21,27]. Even the largest scale analyses, looking at all adult Medicare beneficiaries or the 14 CDC public health tracking states, captured only a quarter of the United States population.

At the time of our study, no articles could be found describing the burden of heat-related emergency conditions in the United States on an annual and national scale. This study aimed to characterize the national trends in ED visits for heat-related emergency conditions in the United States from 2008 – 2019. Our hypothesis was that the incidence rate per 100,000 US population of heat-related ED visits increased significantly over the past decade.

## Methods

### Data Selection

ED records were sourced from the Nationwide Emergency Department Sample (NEDS), provided by the Agency for Healthcare Research and Quality as a part of the Healthcare Cost and Utilization Project [28]. The records provided by NEDS are de-identified and publicly available, so IRB approval was not required for this study. NEDS compiles information from both the State Inpatient Databases and State Emergency Department Databases, capturing patients that are seen in the ED and subsequently admitted to the hospital, transferred to another facility, or released following ED treatment. Data were available from NEDS for the years 2006 – 2019, but this analysis was restricted to 2008 – 2019 due to technical challenges in releasing the 2006 – 2007 NEDS dataset by the Agency for Healthcare Research and Quality. NEDS records were collected from 989 facilities that represent a 20-percent stratified sample of United States hospital-owned EDs [29].

ED visits were included in this study if the medical diagnosis contained an International Classification of Diseases, Ninth Revision Clinical Modifications (ICD-9-CM) or Tenth Revision Clinical Modifications (ICD-10-CM) code for heat-related emergency conditions, as established by previous studies [30]. In the NEDS, ICD-9-CM codes were used from the start of 2008 through quarter 3 of 2015, and included diagnostic codes 992.0-992.9, and external cause of injury codes E9000 and E9009. ICD-10-CM codes were used from quarter 4 of 2015 through the end of 2019, and included codes T67.0-67.9, V932, X30, and X32. Diagnostic codes for accidents or exposure related to excessive man-made heat (E9001 and W92 respectively) were not included, as this analysis was focused on natural heat related emergency conditions.

Weighted counts of heat-related ED visits were generated using the DISCWT method,[28] and estimate the total counts for heat-related ED visits across the United States. A total of 1,007,134 weighted heat-related ED visits were included in this study.

National temperature data were also collected for this study, to allow for a better understanding of the relationship between temperature and heat-related ED visits. Temperature data were sourced from the National Oceanic and Atmospheric Administration Climate at a Glance [31]. This data set compiles daily temperatures taken from each of the 344 climate divisions of the contiguous United States, and weighs divisional values by area to construct national averages [32]. For this analysis, data were gathered on an annual basis, as well as for the months of January, March, and July. January and July were chosen to evaluate the temperature trends in the coldest and hottest months of the year. March was subsequently included due to unanticipated results generated during data analysis for this month.

### Study Variables

For heat-related ED visits included in this study, the date of the visit, hospital geographic location, patient sex, and patient age were analyzed. In this analysis, date of visit was recorded on a monthly time scale. Hospital geographic location was provided by NEDS as Northeast, South, Midwest, and West. As further geographic groupings were not possible due to the limited information provided by NEDS, these same stratifications were utilized during this analysis.

Patient age was provided in years. For the purposes of this study, age groups were generated using findings from previous works. Though inconsistent, studies have noted an increased risk among patients 0 – 4 years for heat-related ED visits [21], emergency medical services calls [33], and mortality [34]. Elderly patients have also been found to be more susceptible to heat-related emergency conditions [35]. An increase in heat-related ED visits has been observed in patients over 65 years old [21], and a step-wise increased risk of mortality has been seen in patients over 65, over 75, and over 85 [36]. Further, school aged children and younger adults are seen to carry an increased risk for heat-related emergency conditions due to recreational and occupational exposures [22]. To best capture these findings, age was analyzed in the following groups: 0 – 4 years, 5 – 17 years, 18 – 49 years, 50 – 64 years, 65 – 79 years, and ≥80 years.

### Statistical Analysis

This study aimed to investigate the changes in the national incidence rate for heat-related emergency conditions from 2008 - 2019. Population data were utilized to generate crude incidence rates per 100,000 US population. Population data were sourced from the Centers for Disease Control and Prevention WONDER online data set for Bridged-Race Population Estimates compiled by the National Center for Health Statistics [37]. This data set utilizes census information to estimate the total population of the United States on July 1^st^ of each year and was collected for the years 2008 – 2019 in this study. Percent change calculations were utilized to examine yearly shifts in incidence rates.

Calculations were made on the general framework of [[(incidence rate 2019) – (incidence rate 2018)] / (incidence rate 2018)] x 100. Averages and 95% confidence intervals were generated for both the incidence rates and percent changes calculated across the study period.

This study also aimed to investigate the burden of heat-related ED visits, as stratified by the selected variables. Weighted counts of heat-related ED visits were used to compare the burden between subgroups. Percent change calculations were utilized to examine yearly shifts in weighted counts of heat-related ED visits and were generated in the same method as described above. Averages and 95% confidence intervals were generated for both the weighted counts and percent changes calculated across the study period.

## Results

Table 1 contains the national estimates for heat-related ED visits from 2008 – 2019, by hospital region, patient age, and sex. Over the study period, an estimated 1,007,134 ED visits for heat-related emergency medical conditions were recorded nationally.

**Table 1.**
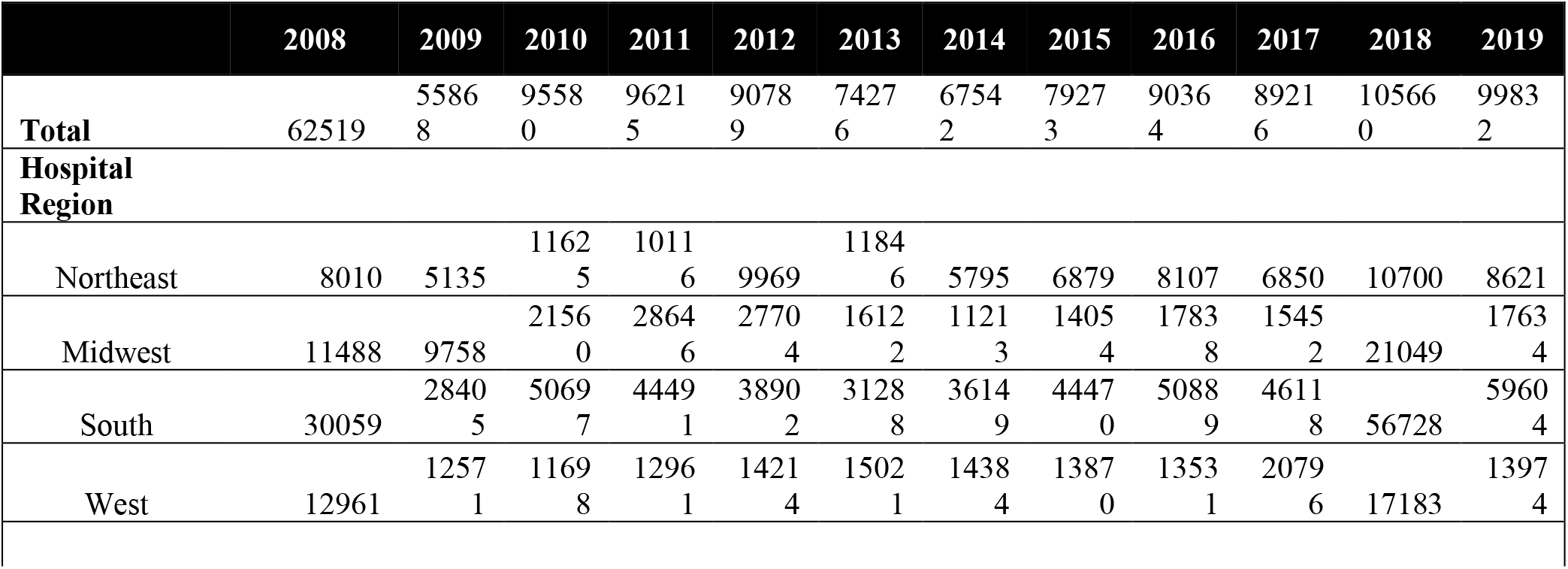
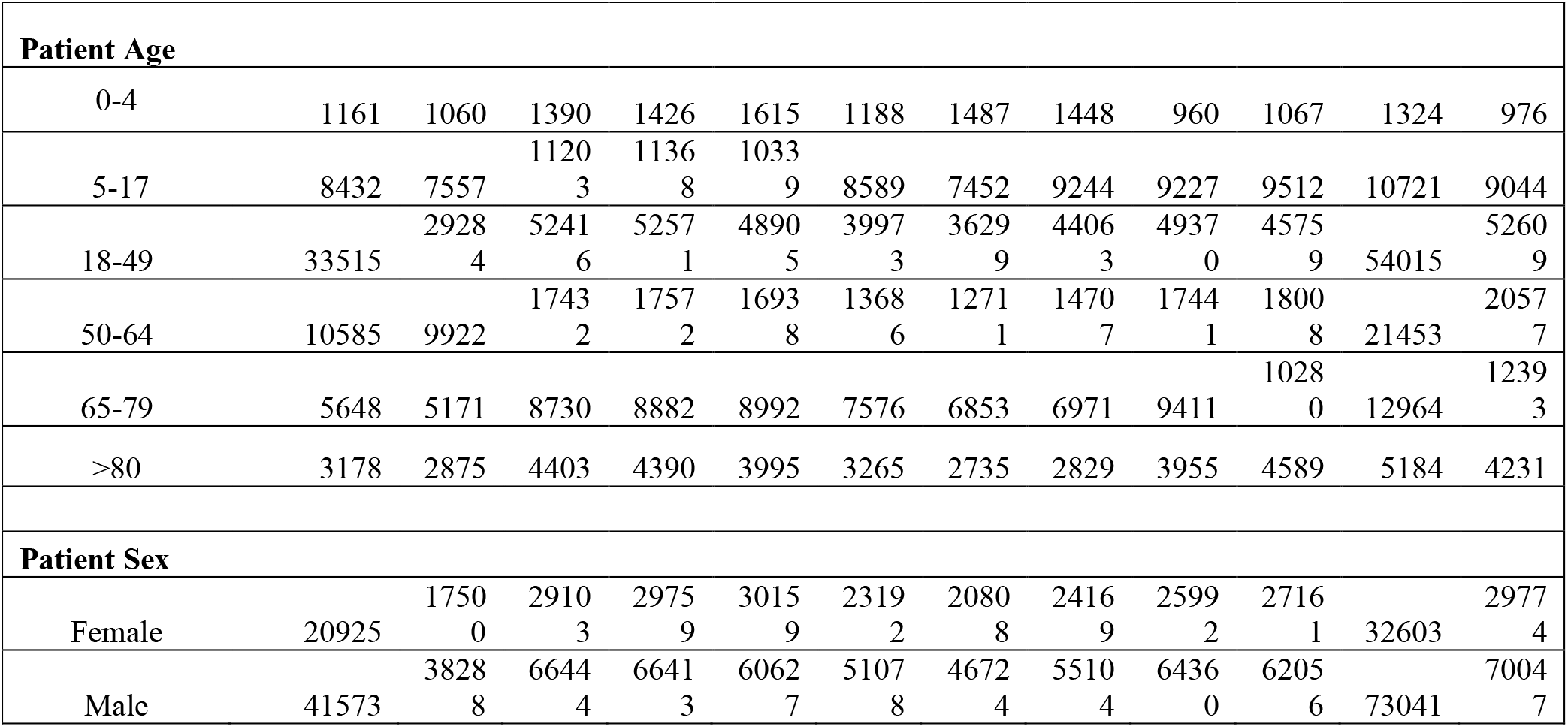
Heat-related ED visit weighted counts, stratified by region, age, and sex, in the United States from 2008-2019.

The South (51.41%) accounted for the most heat-related ED visits over the study period, while the Northeast (10.29%) recorded the least. The Northeast (9.24%, 95%CI: 19.65 – 38.13) and Midwest (11.10%, 95%CI: 15.58 – 37.78) had the greatest average annual increase in heat-related ED visits from 2008 – 2019. Patients aged 0 – 4-years (1.50%) and 5-17 years (11.19%) accounted for a lower percentage of heat-related ED visits than their population percentage (6.3% and 16.95% respectively). Patients aged 18-49 years accounted for the most heat-related ED visits (53.50%), which represented a greater share than their population percentage (43.05%). Patients aged 50 – 64 years (18.97%), 65 – 79 years (10.31%), and ≥ 80 years (4.53%) all accounted for heat-related ED visit percentages similar to their population percentage (19.29%, 10.67%, and 3.74% respectively). Males (69.10%) accounted for the majority of heat-related ED visits during the study period.

Figure 1 displays the annual incidence rate of ED visits for heat-related emergency conditions per 100,000 US population from 2008 - 2019, with the corresponding average temperature across the same time period. Across the study period, the incidence rate increased by 47.9%, from 20.56 in 2008 to 30.41 in 2019 per 100,000 population. The highest incidence rate was observed in 2018 (32.34 per 100,000), and the lowest incidence rate in 2009 (18.21 per 100,000). The average annual incidence rate for the study period was 26.43 (95% CI: 23.78 – 29.08) per 100,000 and increased by an average of 5.73% (95% CI: 8.64% – 20.1%) per year. The greatest increase in incidence rate (69.69%) was observed between 2009 (18.21 per 100,000) and 2010 (30.9 per 100,000). The largest decrease in incidence rate (−18.74%) was seen between 2012 (28.93 per 100,000) and 2013 (23.51 per 100,000). The average annual temperature across the study period was 53.43^0^F (95% CI: 52.82 – 54.04), with the highest temperatures occurring in 2012 (55.28^0^F), and the lowest in 2008 (52.29^0^F). For the months of January, March, and July, the highest average temperatures were all observed in 2012 (36.12^0^F, 50.41^0^F, and 76.77^0^F respectively).

**Figure 1.**
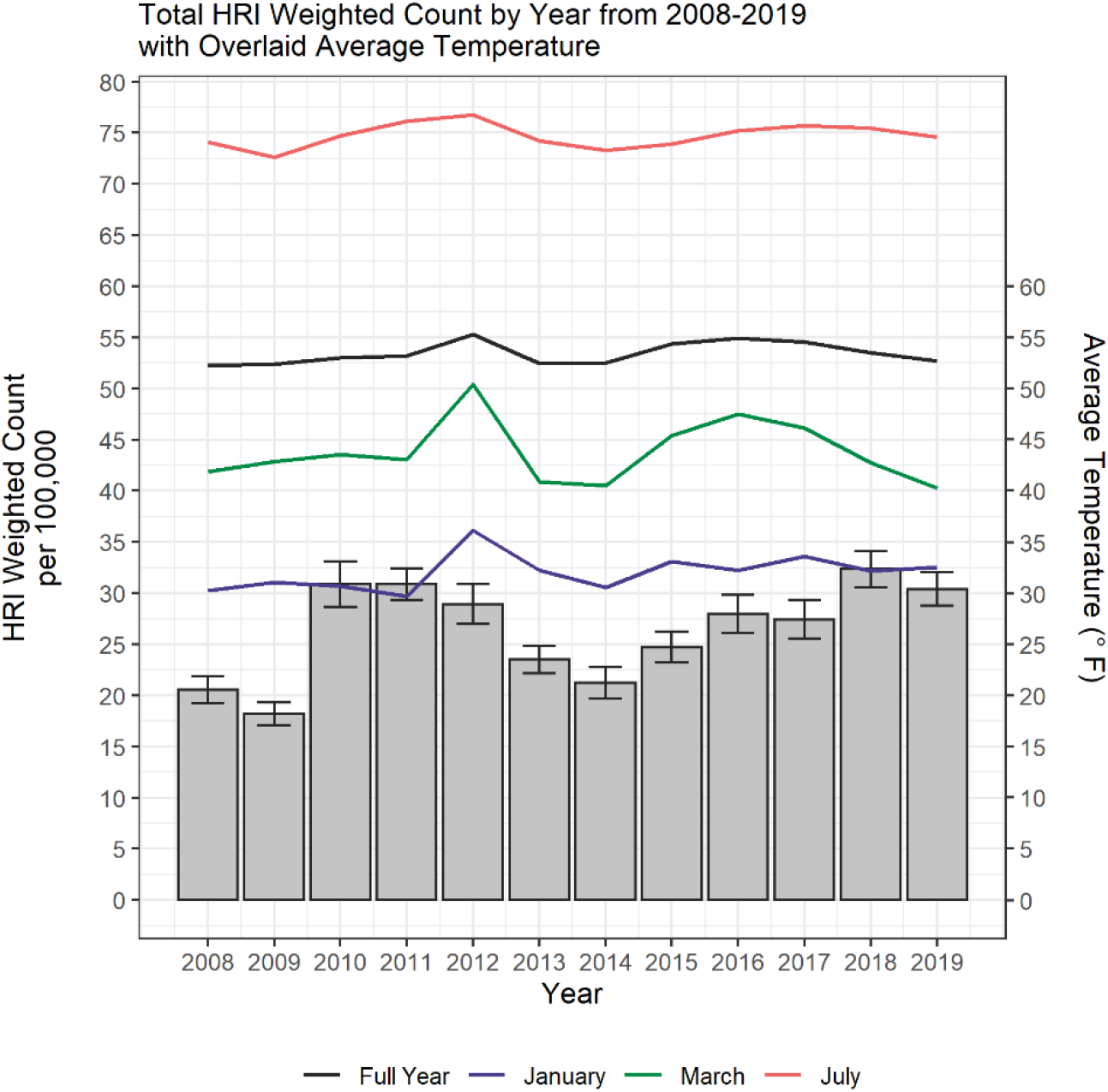
Total HRI weighted count by year from 2008-2019 with overlaid average temperature

Figure 2 depicts the weighted counts for heat-related ED visits stratified by month of visit, for the years 2008, 2012, 2016, and 2019. These years were selected to observe changes in heat-related ED visits at even intervals across the study period. The warm season of May through September accounted for an average of 80.83% (95%CI: 78.69% – 82.97%) of cases annually across the study period. July (29.56%) had the largest case volume over the study period, while December (0.32%) had the smallest. Across the study period, there was an average annual increase of 6.47% (95% CI -8.03% – 20.97%) per year in weighted counts of heat-related ED visits. July (19.25%, 95% CI: 20.75% – 59.26%) and March (14.36%, 95% CI: 19.53% – 48.25%) had the greatest average monthly increases. February (−1.63%, 95% CI: 11.51% – 8.25%) was the only month with an average decrease in weighted counts of heat-related ED visits.

**Figure 2.**
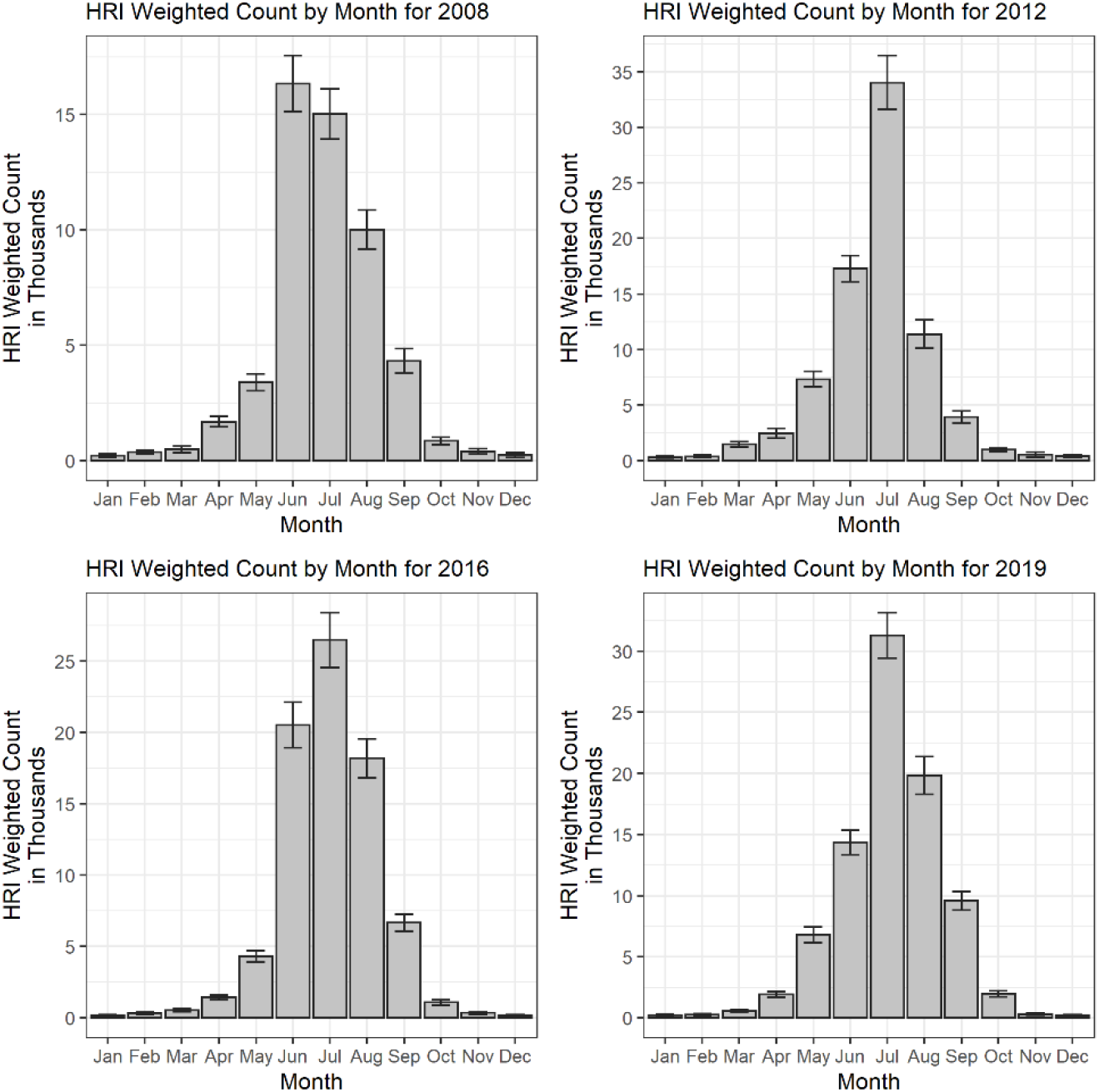
HRI weighted counts by month for 2008, 2012, 2016, 2019

## Discussion

This study, based on national medical records data, explored the trends in ED visits for heat-related emergency conditions across the United States from 2008 to 2019. An overall increase in the incidence rate for heat-related ED visits was observed during the study period, with this increase holding true both nationally and regionally. The burden of heat-related emergency conditions was seen to be greatest in the South. Most cases occurred during the month of July, with case numbers rising at the greatest rate in July and March.

While previous research into heat-related emergency conditions has largely been restricted to narrow populations of interest, the national approach of this study has allowed for a greater assessment of national trends and regional variations in heat-related ED visits across the United States. It is unsurprising that the South accounted for the greatest number of heat-related ED visits, knowing the regional temperature patterns of the United States. Looking at the demographic make-up of the South, this finding agrees with previous literature suggesting an increased risk of heat-related emergency medical conditions in more rural, non-White areas [23,38]. The observed increase in burden of heat-related ED visits is a particular cause for concern in the South, given the region’s workforce. In the coming years, Southern states are expected to carry the greatest occupational risk to heat-related emergency conditions, and would most directly benefit from policies regulating workloads and shift times [39].

While the burden of heat-related ED visits was greatest in the South, the Northeast and Midwest were seen to have the greatest degree of increase in heat-related ED visits across the study period. This observed increase has several plausible explanations. On the basis of climate change, the Northeast has experienced greater warming and higher heat susceptibility as compared to the Southeast [40,41]. From a regional climate perspective, populations living in areas with significant seasonal weather variability are generally less able to adapt to higher temperatures. This is thought to be due to both physiology and the lesser availability of social adaptations, such as air conditioning [15,42]. Thus, focusing interventions on creating more social adaptations – particularly equipping public spaces with air conditioning and increasing tree cover – would be most effective in these regions [43].

When designing interventions for heat-related emergency conditions, it is important to note that the temperature threshold at which susceptibility increases is not the same across regions. Areas with lower average temperature have an increased risk for heat-related emergency conditions at lower temperature thresholds than areas of consistently warm climate [12]. While heat advisory alert systems have different temperature trigger points in different regions of the United States, past studies suggest the current models are not sufficient to both mitigate health risks and limit alarm fatigue [44]. Further research comparing heat advisory systems in different regions is warranted.

Results of this study also suggest that heat advisory trigger points should be evaluated during different months of the year. The burden pattern observed for the month of March suggest heat-related emergency medical risk increases with uncharacteristically high temperatures that are not exclusively limited to the warm season. A large peak of heat-related ED visits were observed in March 2012, which corresponds to the March 2012 heatwave [45]. Record breaking temperatures were recorded across the country, and the national monthly average temperature deviating 8.9^0^F above the twentieth century average [31]. Additional rises in heat-related ED visits were observed in March of 2015 and 2017, which similarly correspond to elevated temperatures in the western regions of the United States during these years [46,47]. As heat waves are predicted to begin earlier in the year [4], higher burdens of heat-related emergency conditions during March of subsequent years can be expected. This poses a particular concern, as extreme heat events in the early months of the year have been linked to greater health risks [48,49]. This is thought to be explained by the body’s lesser ability to adapt to higher temperatures following the winter months. Targeted interventions to limit heat-related emergency conditions enacted during the Spring could therefore be beneficial in the coming years.

### Limitations

Results of this study should be interpreted in the context of limitations. First, the methods used to identify heat-related ED visits did not allow for the distinguishment between primary, subsequent, or sequalae visits (within the ICD-9-CM codes). Thus, the counts reflected in this analysis may inaccurately overestimate the true count of unique patients experiencing heat-related emergency conditions within the population. However, given the course of illness and effectiveness of treatment for heat-related emergency conditions, it is reasonable to assume that the majority of included heat-related ED visits were primary visits caused by heat-related emergency conditions. Second, temperature data examined in this study was collected as a national average. This prevented the exploration into regional temperature patterns and gave rise to the potential for elevated temperatures in certain sectors of the United States to be obscured by depressed temperatures in other areas. Specific heat waves were also unable to be accounted for with this data. Finally, the nature of the ED data obtained from the Healthcare Cost and Utilization Project’s NEDS datasets prevented further patient information from being elicited. For example, it is therefore unknown if the experienced heat-related emergency medical condition was related to occupational or recreational activities. Patient socioeconomic status and race was also left out of analysis due to the availability of patient level data in NEDS for the study period, despite previous works suggesting an association between these variables and heat-related ED visits.

## Conclusion

In this nationwide study, a significant increase in ED visits for heat-related emergency conditions was seen across the United States in the last decade, affecting populations of all ages, in all regions, and during all seasons. As the impact of climate change is predicted to worsen in the coming years, targeted interventions are necessary to mitigate the human health impact of this growing public health crisis.

## Data Availability

All data produced are available online from the Healthcare Cost and Utilization Project website.

https://www.hcup-us.ahrq.gov/nedsoverview.jsp

